# SARS-CoV-2 genomic surveillance identifies naturally occurring truncations of ORF7a that limit immune suppression

**DOI:** 10.1101/2021.02.22.21252253

**Authors:** Artem Nemudryi, Anna Nemudraia, Tanner Wiegand, Joseph Nichols, Deann T Snyder, Jodi F Hedges, Calvin Cicha, Helen Lee, Karl K Vanderwood, Diane Bimczok, Mark Jutila, Blake Wiedenheft

**Affiliations:** Department of Microbiology and Immunology, Montana State University, Bozeman, MT 59717, USA; Gallatin City-County Health Department, Bozeman, MT, 59715, USA

**Keywords:** SARS-CoV-2, ORF7a, IFN response

## Abstract

Over 200,000 whole genome sequences of SARS-CoV-2 have been determined for viruses isolated from around the world. These sequences have been critical for understanding the spread and evolution of SARS-CoV-2. Using global phylogenomics, we show that mutations frequently occur in the C-terminal end of ORF7a. We have isolated one of these mutant viruses from a patient sample and used viral challenge experiments to demonstrate that Δ115 mutation results in a growth defect. ORF7a has been implicated in immune modulation, and we show that the C-terminal truncation results in distinct changes in interferon stimulated gene expression. Collectively, this work indicates that ORF7a mutations occur frequently and that these changes affect viral mechanisms responsible for suppressing the immune response.

**Highlights:**

- ORF7a mutations are found in SARS-CoV-2 genomes isolated from around the globe.
- ORF7a mutation results in a replication defect.
- An ORF7a mutation limits viral suppression of the interferon response.

## Introduction

The spillover of Severe Acute Respiratory Syndrome Coronavirus 2 (SARS-CoV-2) into the human population has resulted in a global pandemic of Coronavirus Disease 2019 (COVID-19) with more than 2.1 million deaths worldwide (https://coronavirus.jhu.edu/) (Andersen et al., 2020). While much of the research on this virus is focused on the Spike protein, recent reports demonstrate that accessory proteins of SARS-CoV-2 might be involved in COVID-19 pathogenesis by modulating antiviral host responses (Young et al., 2020; Zhang et al., 2020). SARS-CoV-2 uses multiple strategies to evade host immunity. Accessory proteins ORF3b, ORF6 and ORF7a antagonize various steps of type I interferon (IFN-I) production and signaling, while ORF8 disrupts antigen presentation by downregulating MHC-I (Konno et al., 2020; Lei et al., 2020; Miorin et al., 2020; Xia et al., 2020; Zhang et al., 2020). To occlude signal transmission from IFN receptors, ORF7a subverts phosphorylation of STAT2, suppressing transcriptional activation of antiviral IFN-stimulated genes (ISGs) that can otherwise restrict viral replication (Martin-Sancho et al., 2020; Xia et al., 2020). While the proposed function for ORF7a is intracellular, antibodies against ORF7a are elevated in the serum of COVID-19 patients (Hachim et al., 2020). Work that is currently under review indicates that recombinant ORF7a protein interacts with CD14^+^ monocytes and triggers expression of pro-inflammatory cytokines, including interleukin-6 (IL-6) and tumor necrosis factor alpha (TNF-*α*) (Zhou, Z. et al., 2020). However, it is unclear if these interactions happen in COVID-19 patients. Together, these data suggest that ORF7a plays a dual role in SARS-CoV-2 infection by modulating both the IFN and inflammatory responses.

Here, we show that C-terminal mutations in ORF7a occur frequently in samples isolated from patients around the globe. These mutations are not derived from a single lineage and they do not persist over time. Using samples collected from infected patients, we have isolated a virus containing a deletion mutation in ORF7a that truncates the C-terminal half of the protein. In vitro viral challenge experiments demonstrate that this mutation results in a replication defect and obviates viral suppression of the immune response. Collectively, these data suggests that ORF7a truncations are defective in suppressing the host immune response, which may explain why these mutations quickly disappear in the immunocompetent population.

## Results

### SARS-CoV-2 genomic surveillance identifies truncation of ORF7a accessory protein

Genome sequencing has been used to track the rise and spread of new SARS-CoV-2 lineages over the course of the pandemic (https://www.gisaid.org/) (Bedford et al., 2020; Korber et al., 2020). As part of this effort, we sequenced SARS-CoV-2 genomes isolated from patients in Bozeman, Montana (Figure 1A, Table S1). In total, we determined 56 whole genome sequences of SARS-CoV-2 viruses isolated from patients between April and July of 2020 using an amplicon-based Nanopore protocol from the ARTIC network (https://artic.network/) (Quick et al., 2017; Tyson et al., 2020). To place the local outbreak in the context of the global SARS-CoV-2 evolutionary trends, we aligned 56 whole genome sequences isolated from patients in Bozeman to 4,235 genomes subsampled (10 genomes per month per country) from 181,003 SARS-CoV-2 genomes downloaded from the GISAID. The subsampling protocol removes redundancy and bias introduced by uneven distribution of the global SARS-CoV-2 sequencing effort. The resulting alignment was used to build a phylogenetic tree (Figure 1B). Of the 56 SARS-CoV-2 genomes determined from patients in Bozeman, only one was derived from the WA1 lineage (lineage A in Figure 1B) that was introduced to Washington state from Wuhan, China in January 2020 (Bedford et al., 2020; Holshue et al., 2020). The remaining 55 genomes associate with clade B.1 and its offshoots, which are characterized by the D614G mutation in the spike and have prevailed globally since the spring of 2020 (Rambaut, Andrew et al., 2020)(Figure 1B). The genetic variability in SARS-CoV-2 circulating in Bozeman (April to July 2020) represents fourteen independent viral lineages, reflecting multiple introductions of the virus to the community from many different sources (Figure S1A).

**Figure 1.**
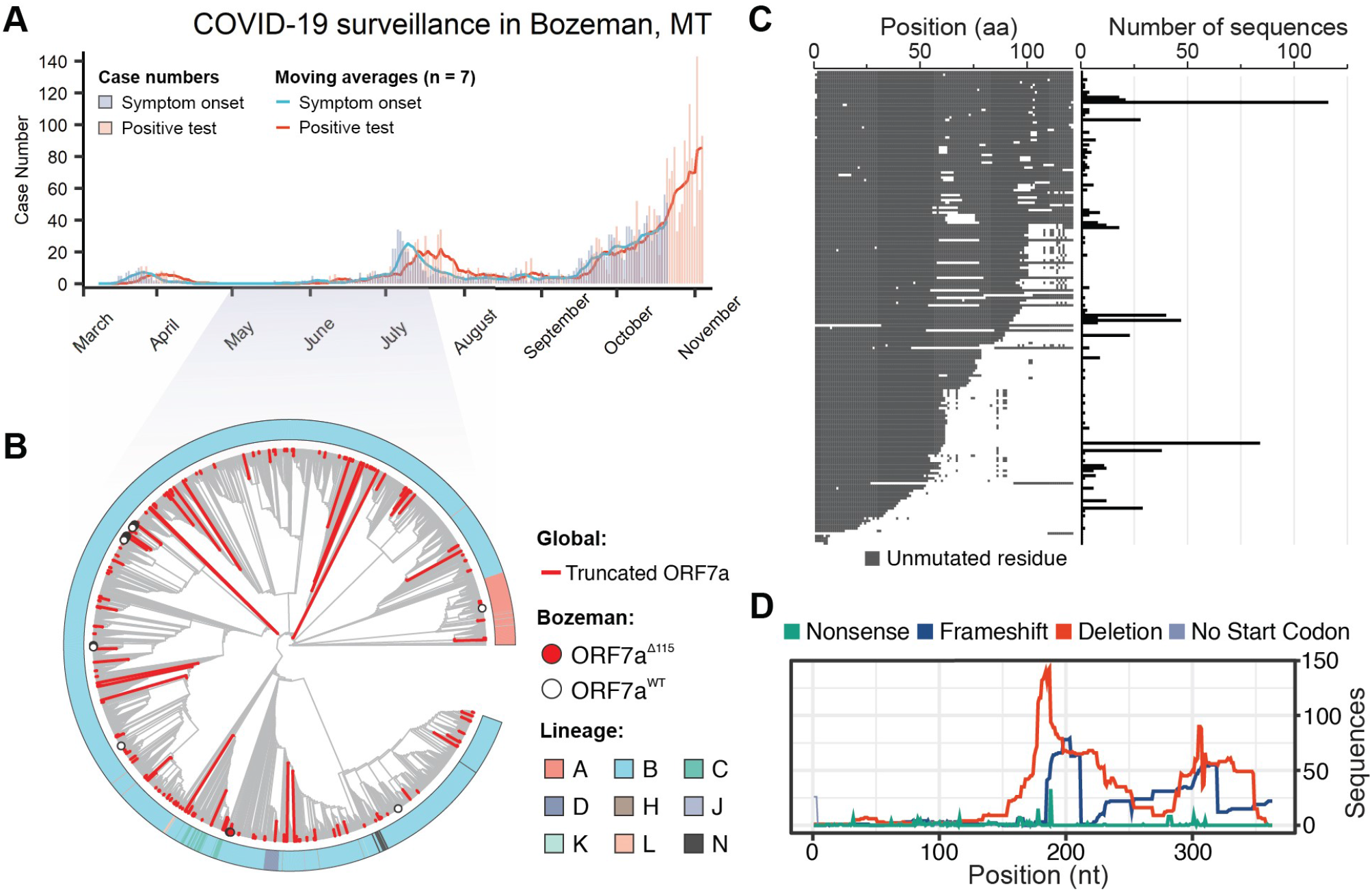
SARS-CoV-2 genomic surveillance identifies global reoccurrence of ORF7a truncations. **A)** Symptom onset (purple) and PCR-based SARS-CoV-2 test results (coral) for patients in Bozeman, Montana are shown with vertical bars. Seven day moving averages, shown with lines, were used to indicate epidemiological trends. **B)** Phylogenetic analysis of SARS-CoV-2 genomes sampled in Bozeman and globally. The tree was constructed from an alignment of 56 Bozeman samples and 4,871 genomes subsampled from GISAID. Subsampling was performed using Augur utility (Nextstrain) by selecting 10 genomes per country per month since the start of the pandemic. Red branches identify truncated ORF7a variants (n= 205) detected in the global data and merged into the alignment. The red dot highlights seven of the 56 ORF7a variants that were isolated in Bozeman between April and July (2020). White dots highlight 49 viral genomes isolated in Bozeman that have wild-type ORF7a sequences. **C)** Alignment of unique ORF7a protein sequences with five or more mutations as compared to the SARS-CoV-2 reference strain (Wuhan-Hu-1). Conserved residues are depicted in gray, while mutations and deletions are shown in white. The frequency of each genotype is shown on the right. **D)** Distribution of different mutations that occur along the ORF7a coding sequence.

During the annotation of these genomes, we noticed a reoccurring (7 out of 56 genomes) 115 nt deletion in the gene encoding accessory protein ORF7a (27,549 - 27,644 nt). The ORF7a^Δ115^ mutation was found in patient swabs collected over a period of 1.5 months (Table S1). RT-PCR and Sanger sequencing verified that all these seven ORF7a^Δ115^ variants are bona fide mutations and not a sequencing artifact (Figure S1B, C). In addition to the ORF7a^Δ115^ deletion, these genomes also share 10 single nucleotide variants (SNVs) compared to the Wuhan-Hu-1 reference genome. Seven of the 10 SNVs are found frequently worldwide and are a signature of the B1.1 lineage of SARS-CoV-2 (Rambaut, Andrew et al., 2020). The remaining 3 mutations are rare, do not co-occur in any other genomes on GISAID, and lead to amino acid changes in ORF3a (Q38P, L95F) and N (R195I) proteins. Interestingly, one of the genomes in the ORF7a^Δ115^ cluster has two additional SNVs that likely were acquired during circulation in the community. This virus was sampled from a patient 41 days after sampling the first ORF7a^Δ115^ variant, which agrees well with estimated SARS-CoV-2 mutation rates (2 nucleotides / month) (van Dorp et al., 2020).

To determine if the ORF7a^Δ115^ genotype is unique to Bozeman we downloaded an alignment of 180,971 SARS-CoV-2 genomes from GISAID, extracted ORF7a sequences and determined their mutational profiles. In total, we identified 845 unique ORF7a gene variants that are different from the Wuhan-Hu-1 reference sequence. Next, we looked for mutations with major effect on the ORF7a amino acid sequence. We identified 189 unique ORF7a variants (825 total sequences) with frameshifts, deletions or missense mutations causing premature stop codons (Figure 1C). To understand the evolutionary history of these mutations, we integrated these genome sequence into our phylogenetic analysis. Detected ORF7a variants appear to have emerged independently on every continent and were not confined to any single lineage (Figure 1B; Table S1). Next, we aligned translated ORF7a sequences to look for patterns. Most of the identified ORF7a mutations (126 out of 189) truncate the C terminus but preserve the N-terminal half of the protein (Figure 1C and D). A portion (36.5%) of these truncated ORF7a variants appeared in two or more patient samples (116 max). While some of these ORF7a mutants appear to have arisen on multiple independent occasions, others come from genomes that form monophyletic clades, suggesting that viruses with ORF7a truncations are capable of transmission within a host population (Figure 1C).

### ORF7a truncation changes protein localization

ORF7a is a type I transmembrane protein with an N-terminal immunoglobulin-like (Ig-like) ectodomain, stalk, transmembrane (TM) domain and a short cytosolic tail (Figure 2A). The two *β* sheets of the ectodomain are held together by two disulfide bonds (i.e., Cys23-Cys58 and Cys35-Cys67) (Figure 2B). The cytosolic tail of ORF7a contains a dilysine (KRKTE) ER retrieval signal (ERRS) that mediates protein trafficking to the ER-Golgi intermediate compartment (ERGIC) (Nelson et al., 2005).

**Figure 2.**
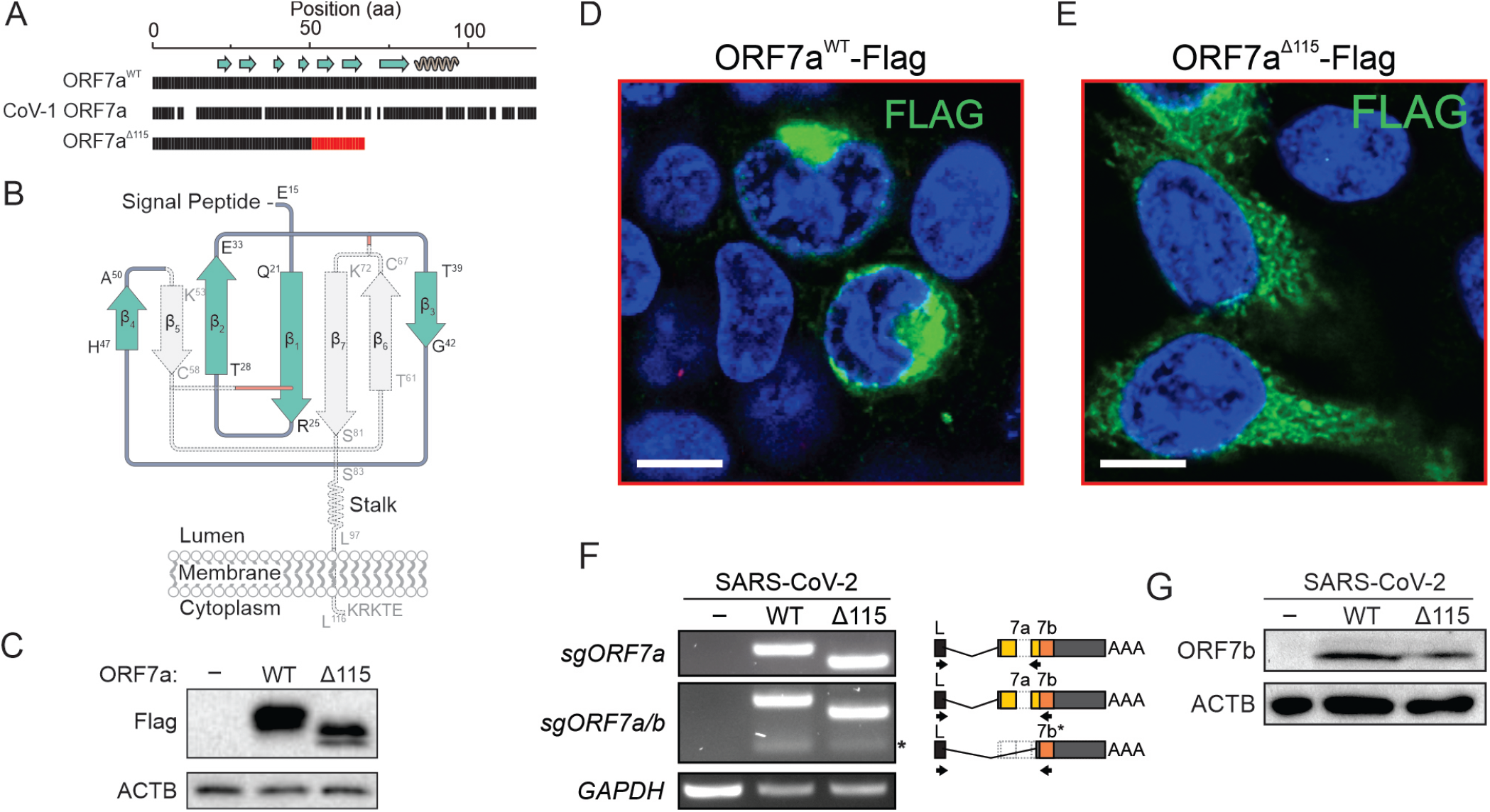
ORF7a truncation results in loss of cellular localization. **A)** Amino-acid (aa) sequence alignment of SARS-CoV-2 ORF7a^WT^, ORF7a^Δ115^ and SARS-CoV-1 ORF7a. Gaps show non-matching positions, red shows 17 aa sequence resulting from a frameshift in the ORF7a mutant. Beta strands (arrows) and alpha helices (coil) are shown above the alignment. Diagram of SARS-CoV-2 ORF7a Ig-like fold. Disulfide bonds that stabilize the *β*-sandwich structure are shown with red lines. The portion of the protein eliminated by the deletion is shown in gray. **C)** C-terminally Flag-tagged ORF7a^WT^ and ORF7a^Δ115^ were cloned and overexpressed in HEK 293T-hACE2. Protein expression was confirmed with Western Blot using anti-Flag antibody. *β*-actin (ACTB) was used as a loading control. **D)** Flag-tagged ORF7a^WT^ or **E)** ORF7a^Δ115^ expressed in HEK 293T-hACE2 cells. Immunostaining preformed using an anti-Flag antibody (green). Cell nuclei were stained with Hoechst 33342 (blue). White scale bar is 10 *µ*m. **F)** ORF7a and ORF7b sgRNAs were identified by RT-PCR. Lower MW band corresponding to ORF7b sgRNA is indicated with asterisk (*). Diagram on the right shows primer (arrows) positions. Specificity of PCR products was confirmed with sanger sequencing. GAPDH was used as a control for cDNA synthesis. **G)** Lysates from SARS-CoV-2 infected cells were probed with antibodies raised against ORF7b protein. ACTB was used as a loading control.

The Δ115 nt mutation in ORF7a introduces a premature stop codon that eliminates *β* 5, *β* 6 and *β* 7, two of the cystines that form disulfides (Cys58 and Cys67), the TM and the cytosolic tail (Figures 2A and B). This truncation is expected to destabilize the protein structure and significantly impact protein function and trafficking in the host cell. To determine how loss of TM and sorting signal affects protein localization, we cloned and expressed Flag-tagged wildtype and truncated ORF7a proteins in HEK 293T-hACE2 cells. The wildtype ORF7a accumulated in the perinuclear region of the cell, which is consistent with the previously reported ERGIC localization (Figure 2D) (Martin-Sancho et al., 2020; Nelson et al., 2005). In contrast, the truncated ORF7a is distributed throughout the cytoplasm and does not associate with specific subcellular compartments, which is consistent with the loss of the TM domain and the ERRS signals required for protein targeting (Figure 2E). Overall, we predict that the extent of the truncation and the loss of intracellular targeting is likely to affect ORF7a function (Gordon et al., 2020; Taylor et al., 2015).

### ORF7a mutation has no collateral effect on ORF7b

The ORF7a gene overlaps with the downstream ORF7b gene (Figure S2). To determine if the Δ115 mutation in ORF7a impacts ORF7b, we first examined whether it eliminates a transcription-regulatory sequence (TRS) that is required for subgenomic RNA (sgRNA) synthesis. None of the TRSs identified via direct RNA sequencing of SARS-CoV-2 overlap with the Δ115 mutation site (Kim et al., 2020), suggesting that ORF7b transcription is not affected (Figure S2). To confirm this prediction, we used RT-PCR with primers that detect ORF7a and ORF7b sgRNAs (Figure 2F). In this assay, we infected 293T-hACE2 cells (MOI = 0.05) with ORF7a^WT^ or ORF7a^Δ115^ viral strains and extracted total RNA from cells at 24 hours post infection (hpi). Both viruses produced specific RT-PCR products corresponding to ORF7a and ORF7b sgR-

NAs. Finally, to verify that the ORF7a^Δ115^ variant does not impact ORF7b translation, we probed cell lysates 24 hpi with an anti-ORF7b antibody (Figure 2G). We detected ORF7b protein in both viral strains with band intensities that suggest similar expression levels. Collectively, these data indicate that the ORF7a^Δ115^ mutation has no collateral effect on the adjacent gene.

### ORF7a truncation results in a replication defect

To determine if ORF7a truncations impact viral replication, we infected 293T-hACE2 and Vero E6 cells with ORF7a^WT^ and ORF7a^Δ115^ viruses at an MOI 0.05 and measured viral RNA levels using RT-qPCR (Figure 3A and B). In this assay, we used viral strains sharing the same haplotype (C241T, C3037T, C14408T, A23403G) that defines the SARS-CoV-2 lineage and its derivatives (Figure S3A) (Rambaut, Andrew et al., 2020). Over the course of 120 hours, ORF7a^WT^ viral RNA level increased 960-fold in supernatants from Vero E6 cells (Figure 3A) and 270-fold in supernatants from 293T-hACE2 cells (Figure 3B). The ORF7a^Δ115^ virus displayed limited replication that resulted in only 18.6-fold and 7-fold RNA level increase at 120 hpi in VeroE6 and 293T-hACE2, respectively. This growth defect reaches statistical significance starting at 6 hpi in Vero E6 cells (p-value = 0.0135) and 24 hpi in 293T-hACE2 cells (p-value = 6.2*10^−5^).

**Figure 3.**
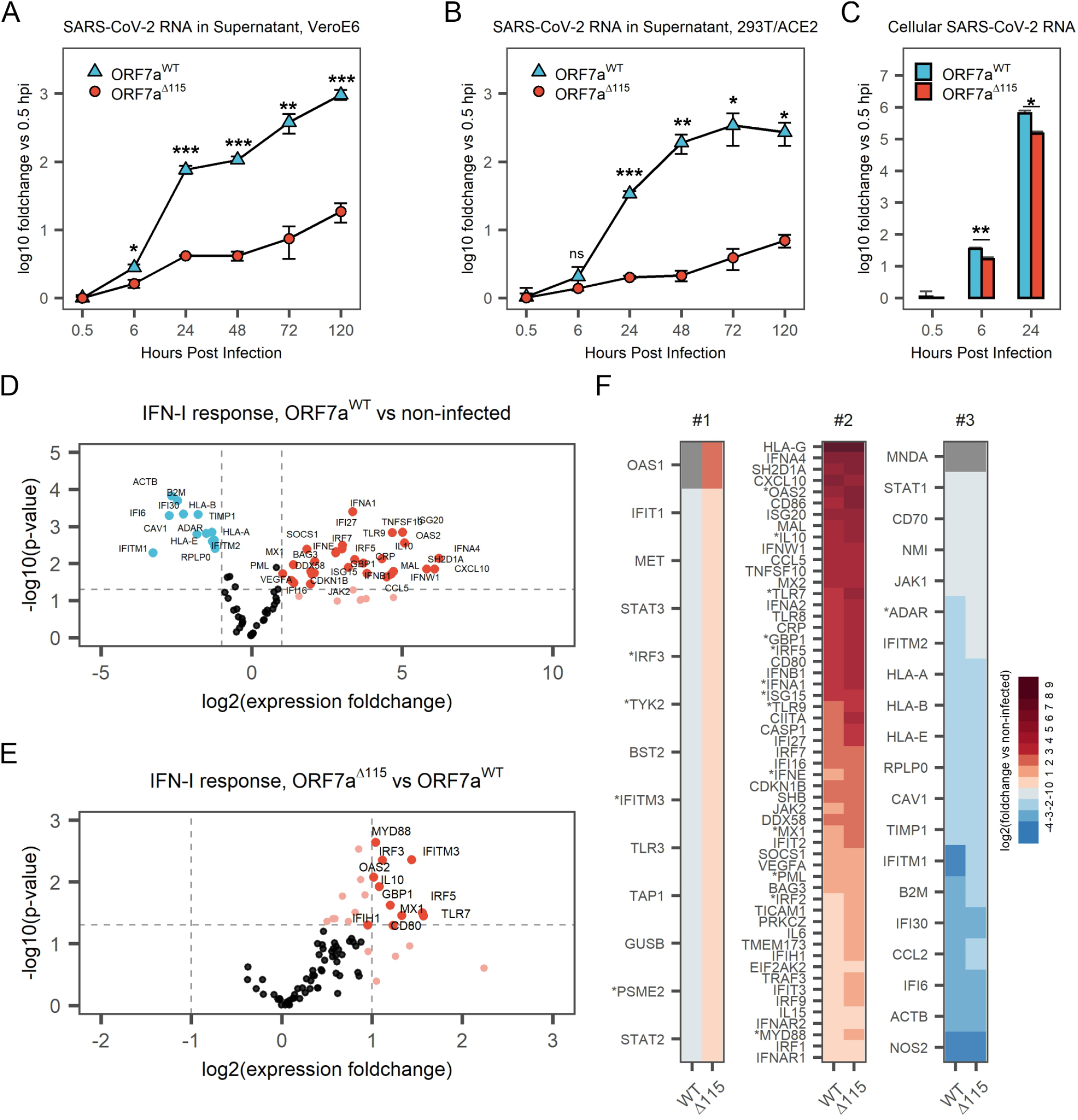
ORF7a truncation results in loss of cellular localization. **A)** Amino-acid (aa) sequence alignment of SARS-CoV-2 ORF7a^WT^, ORF7a^Δ115^ and SARS-CoV-1 ORF7a. Gaps show non-matching positions, red shows 17 aa sequence resulting from a frameshift in the ORF7a mutant. Beta strands (arrows) and alpha helices (coil) are shown above the alignment. **B)** Diagram of SARS-CoV-2 ORF7a Ig-like fold. Disulfide bonds that stabilize the *β*-sandwich structure are shown with red lines. The portion of the protein eliminated by the deletion is shown in gray. **C)** C-terminally Flag-tagged ORF7a^WT^ and ORF7a^Δ115^ were cloned and overexpressed in HEK 293T-hACE2. Protein expression was confirmed with Western Blot using anti-Flag antibody. *β*-actin (ACTB) was used as a loading control. **D)** Flag-tagged ORF7a^WT^ or **E)** ORF7a^Δ115^ expressed in HEK 293T-hACE2 cells. Immunostaining preformed using an anti-Flag antibody (green). Cell nuclei were stained with Hoechst 33342 (blue). White scale bar is 10 *µ*m. **F)** ORF7a and ORF7b sgRNAs were identified by RT-PCR. Lower MW band corresponding to ORF7b sgRNA is indicated with asterisk (*). Diagram on the right shows primer (arrows) positions. Specificity of PCR products was confirmed with sanger sequencing. GAPDH was used as a control for cDNA synthesis. **G)**) Lysates from SARS-CoV-2 infected cells were probed with antibodies raised against ORF7b protein. ACTB was used as a loading control.

To determine if this phenotype results from a defect in viral egress only, or if viral RNA replication in the host is also affected, we examined early steps in the infection. We extracted total RNA from the infected 293T-hACE2 cells at 0.5, 6 and 24 hpi, and measured SARS-CoV-2 RNA with RT-qPCR. Compared to wildtype, ORF7a^Δ115^ RNA level was reduced 2.1-fold inside the host at 6 hpi (p-value = 0.002) and 4.3-fold at 24 hpi (p-value = 0.02) (Figure 3C). This decrease in viral RNA in the host cell differs from the corresponding decrease in the supernatant RNA at 24 hpi (4.3-fold vs 16.8-fold, respectively; Figure 3B, C). This disparity suggests that the ORF7a^Δ115^ mutation might affect genomic RNA replication and transcription, as well as viral egress.

### ORF7a^Δ115^ mutation alters host Type I interferon response to SARS-CoV-2

The ORF7a protein is not essential for SARS-CoV-2 replication (Xia et al., 2020). When overexpressed in vitro, ORF7a suppresses type I IFN (IFN-I) signaling (Xia et al., 2020). Interferon stimulated genes (ISGs) exert anti-viral activities and thus we hypothesized that the replication defect in ORF7a^Δ115^ results from impaired suppression of host anti-viral immune responses. To test this hypothesis, we measured expression of type I IFN response genes upon infection with ORF7a^WT^ and ORF7a^Δ115^ strains using an RT-qPCR array targeting 91 human transcripts (Table S2). The array included 3 reference genes that we used to normalize expression of 88 target genes. In agreement with recent transcriptomic studies, we detected a type I IFN-mediated antiviral response after infecting HEK 293T-hACE2 with SARS-CoV-2 isolates (Figure 3D, Figure S3B) (Banerjee et al., 2020; Blanco-Melo et al., 2020). Out of 88 type I IFN response genes included in the RT-qPCR array, 42 were differentially expressed (*≥* 2-fold change, p-value *<*0.05) in ORF7a^WT^ infected cells and 55 in ORF7a^Δ115^ infection, as compared to the non-infected control (Figure 3D, Figure S3B). Both viruses stimulated expression of multiple type-I interferon genes, antiviral ISGs, interleukins and proinflammatory cytokines (Table S2).

To understand the effect of ORF7a^Δ115^ on the host type I IFN response, we compared expression of genes targeted with RT-qPCR array between the two viruses. In general, the ORF7a^Δ115^ variant induces elevated type I IFN response to the infection (Figure S3C, D). Analysis of genes differentially expressed between the two viral infections produced an asymmetric volcano plot gravitating towards upregulation (Figure 3E). A subset of ISGs had expression levels that are significantly different (p-value *<*0.05) between the two viral infections (Figure 3E, F, Table S2). This subset includes sensors (TLR7), signal transducers (MYD88, OAS2), transcriptional regulators (IRF3, IRF5) and restriction factors (GBP1, IFITM3, MX1) known to combat infections by RNA viruses (Schoggins, 2019). One of these differentially expressed ISGs (IFITM3) restricts SARS-CoV-2 entry (Shi et al., 2020; Zang et al., 2020). Additionally, polymorphisms in IFITM3, MX1 and TLR7 are linked to the severity of COVID-19 (Andolfo et al., 2020; Jin, R. et al., 2020; Pati et al., 2021). This upregulation of ISGs indicates that the ORF7a^Δ115^ mutation limits viral suppression of the IFN-I response, which is consistent with previous work suggesting that ORF7a prevents transcriptional activation by STAT1/STAT2 complex (Xia et al., 2020). Collectively, our data show that a naturally occurring truncation variant of the ORF7a protein alters the host innate immune responses to the in vitro infection.

## Discussion

In this study, we use genomic surveillance and phylogenetics to identify ORF7a variants that have emerged globally throughout the SARS-CoV-2 pandemic. Local occurrence of ORF7a mutations has been reported by others (Addetia et al., 2020; Holland et al., 2020; Rosenthal et al., 2020). However, the effect of these mutations on viral fitness and host immune responses has not been investigated. Here we have isolated an ORF7a^Δ115^ variant and show that this mutation results in an in vitro growth defect. This defect is coupled with elevated IFN response to SARS-CoV-2 and specifically ISGs.

Interferon systems are a frontline of host defense that signals infection and elicits an antiviral state (McNab et al., 2015). Pretreatment with type I, type II and type III IFNs restricts SARS-CoV-2 infection and replication in cell culture (Mantlo et al., 2020; Miorin et al., 2020). SARS-CoV-2 deploys multiple proteins (e.g., nsp6, nsp13, ORF6, ORF7a, ORF7b and ORF9b) to shut down IFN signaling and dampen innate immune responses (Jiang et al., 2020; Lei et al., 2020; Xia et al., 2020). In particular, when overexpressed ORF7a subverts STAT2 phosphorylation, blocking IFN-dependent transcriptional activation of antiviral ISGs (Xia et al., 2020). Both type I and III IFNs signal through the JAK-STAT pathway and overlap substantially in the transcriptional responses they induce (Kotenko and Durbin, 2017). Therefore, ORF7a likely suppresses both IFN types, however its role in suppressing the type III IFN response has yet to be verified (Xia et al., 2020). The ORF7a^Δ115^ variant impairs viral replication in HEK 293T-hACE2 as well as in Vero E6, the latter of which do not express type I IFN genes (Emeny and Morgan, 1979; Osada et al., 2014). While type I IFN genes are deleted in Vero E6 cells, the type III IFNs are intact and are activated upon infection with RNA viruses, which results in ISG upregulation (Stoltz and Klingstrom, 2010; Wang et al., 2020). Taken together, these data suggest that the growth defect of the ORF7a^Δ115^ strain in Vero E6 cells might result from a type III IFN-dependent response that the virus fails to suppress.

Cytokine profiling and RNA-seq of clinical samples reveal suppressed or delayed IFN responses in COVID-19 patients (Arunachalam et al., 2020; Hadjadj et al., 2020; Lucas et al., 2020). Dysregulated immune responses coupled with exacerbated inflammatory cytokines drive pathogenesis of the disease and correlate with its severity (Jin, X. et al., 2020; Tay et al., 2020; Zhou, F. et al., 2020). Here, we detected enhanced IFN signaling in cells infected with a naturally occurring ORF7a^Δ115^ variant of SARS-CoV-2. We anticipate that major truncation and loss of protein targeting (Figure 2) limit the ability of ORF7a to suppress IFN signaling. Given the importance of the IFN response in COVID-19, we anticipate that ORF7a mutations might affect SARS-CoV-2 pathogenicity. Clinical comparisons will be required to examine this potential effect on COVID-19 features.

Deletions in ORF7a, 7b and ORF8 have previously been reported (Addetia et al., 2020; Holland et al., 2020; Rosenthal et al., 2020; Su et al., 2020). Similar deletions emerged in MERS- and SARS-CoV, which were linked to viral attenuation (Chinese, 2004; Muth et al., 2018). A 382-nt deletion in ORF8 is associated with milder SARS-CoV-2 infection in patients (Young et al., 2020). While this mutation affects clinical features of COVID-19, it has no evident effect on viral replication or host transcriptional responses in human cell culture (Pekosz et al., 2020). Overexpression of ORF8 results in post-translational downregulation of MHC-I, therefore the protein might modulate the interaction between infected cells and immune system (Zhang et al., 2020).

A new lineage of SARS-CoV-2 (i.e., B.1.1.7) has recently emerged (Rambaut, Andrew et al., 2020). Aside from changes in the Spike protein, this lineage also includes a premature stop codon in ORF8 that truncates most of the protein (Volz et al., 2021). It has been hypothesized that B.1.1.7 might originate from an immunocompromised patient with chronic SARS-CoV-2 infection treated with convalescent plasma (Baang et al., 2021; Bazykin et al., 2021; Rambaut, Andrew et al., 2020). Several studies illustrate how persistent SARS-CoV-2 infections associate with accelerated viral evolution (Avanzato et al., 2020; Choi et al., 2020). In immunocompromised hosts the selective pressure on anti-immune accessory genes is lifted, which might permit loss-of-function mutations. However, these mutations will not persist in the immunocompetent population (Young et al., 2020), unless they co-occur with mutations that increase virus transmissibility or become fixed due to the founder effect (Lauring and Hodcroft, 2021; Rambaut, Andrew et al., 2020).

### Limitations of Study

The viral replication defect and cellular immune responses presented here were performed using in vitro cell culture. While HEK 293T-hACE2 and Vero E6 cells support virus entry and replication (Cantuti-Castelvetri et al., 2020; Chu et al., 2020), they may not recapitulate particular aspects of an in vivo infection (Takayama, 2020). We anticipate that the in vitro results presented here will provide important context for clinical comparisons, similar to those recently published for ORF8 (Young et al., 2020).

## Materials and Methods

### EXPERIMENTAL MODEL AND SUBJECT DETAILS

#### Cell cultures

HEK 293T-hACE2 cells (BEI, NR-52511) were generously provided by Dr. Seth Pincus (Montana State University) and maintained at 37C and 5% CO2 in Dulbecco’s Modified Eagle Medium (DMEM, Gibco, cat. #12100-061) supplemented with 10% fetal bovine serum (FBS, ATLAS Biologicals, Lot. #F31E18D1), sodium bicarbonate (3.7 g/L), 50 I.U./mL penicillin and 50 *µ*g/mL streptomycin. Vero E6 cells (ATCC ® cat. #CRL-1586) were maintained at 37C and 5% CO2 in Eagle’s Modified Eagle Medium (EMEM, ATCC ®, cat. #30-2003) supplemented with 10% fetal bovine serum (FBS, ATLAS Biologicals, Lot. #F31E18D1) and 50 I.U./mL penicillin and 50 *µ*g/mL streptomycin. All cell lines tested negative for mycoplasma.

#### Plasmids

Human-codon optimized dsDNA gene fragments (gBlocks) encoding for SARS-CoV-2 C-terminally FLAG-tagged ORF7aWT and ORF7a^Δ115^ proteins were synthesized by Integrated DNA Technologies (IDT). Sequence of ORF7a from Wuhan-Hu-1 reference (genbank MN908947.3) was used for ORF7aWT gBlock. The gene fragments were cloned into the pEGFP-N1 (Clontech cat# 6085-1) backbone to replace the eGFP gene.

#### Antibodies

Western blotting: Rabbit anti-ACTB (Cat: AC026, 1:20,000) antibodies were from ABclonal, Mouse anti-FLAG antibodies (Cat: MA1-91878-1MG, 1:1000) were from ThermoFisher Scientific, sheep anti-ORF7b antibodies (1:120) were from MRC I PPU, College of Life Sciences, University of Dundee, Scotland; Goat anti-mouse IgG peroxidase-conjugated (Cat: 115-035-003, 1:10,000), Goat anti-rabbit IgG peroxidase-conjugated (Cat: 111-035-003, 1:10,000) and Donkey Anti-Sheep IgG peroxidase-conjugated (Cat: 713-035-147, 1:10,000) antibodies were from Jackson ImmunoResearch. Immunocytochemistry: Mouse anti-FLAG antibodies (Cat: MA1-91878-1MG, 1:200), Goat anti-mouse-AlexaFluor-488 (Cat: A11001, 1:2,000) antibodies were from Invitrogen.

#### Human clinical sample collection and preparation

Clinical samples were obtained with local IRB approval (protocol #DB033020) and informed consent from patients undergoing testing for SARS-CoV-2 at Bozeman Health Deaconess Hospital. Nasopharyngeal swabs from patients that tested positive for SARS-CoV-2 were collected in viral transport media. RNA was extracted from all patient samples using QIAamp Viral RNA Mini Kit (QIAGEN) a biosafety level 3 (BSL3) laboratory. All samples were heat-inactivated before removing from BSL3.

#### Production and titration of coronavirus stocks

The nasopharyngeal swabs in the viral transport media were used to generate viral stocks as previously described (Harcourt et al., 2020). Briefly, 100 ul of the viral transport media was two-fold serially diluted and applied to Vero E6 cells When infected cells showed extensive cytopathic effect, the media was collected and utilized to generate a greater volume second passage in Vero E6 cells. When CPE was apparent, supernatants were centrifuged 1,000 RCF for 5 minutes to remove cellular debris. Virus titer in clarified supernatants was determined with plaque assay in Vero E6 cells as described(Loveday et al., 2020). For plaque assay, Vero E6 cells were incubated with viral inoculum at limiting dilutions. Inoculated cells were overlayed with either 0.75% methylcellulose, DMEM supplemented with 2% FBS and 1% penstrep and incubated for 4 days. Cells were fixed and stained with 0.5% methylene blue in 70% ethanol. Plaques were counted and the overall titer was calculated. Viral stocks identity was confirmed via whole genome sequencing on Oxford Nanopore. All SARS-CoV-2 experiments were performed in a BSL3 laboratory.

#### Symptom onset data and clinical test results

Suspect cases of COVID-19 were tested in a CLIA lab and instructed to self-quarantine until notified of the RT-qPCR test results. All laboratory confirmed positive cases of COVID-19 were contacted via telephone by local public health nurses to complete contact tracing. During this interview, the nurses collected recorded symptoms, symptom onset date, travel history, contact with other known laboratory confirmed cases, close contacts and activities on the two days before symptom onset up until notification of a positive test. Data collection was conducted as part of a public health response. The study was reviewed by the Montana State University Institutional Review Board (IRB) For the Protection of Human Subjects (FWA 00000165) and was exempt from IRB oversight in accordance with Code of Federal regulations, Part 46, section 101. All necessary patient/participant consent has been obtained and the appropriate institutional forms have been archived.

### METHOD DETAILS

#### Reverse Transcription quantitative PCR (RT-qPCR)

RT-qPCR was performed using CDC primers (N1 and N2) and probes from the 2019-nCoV RUO Kit (IDT# 10006713). SARS-CoV-2 RNA was quantified using one-step RT-qPCR in ABI 7500 Fast Real-Time PCR System according to CDC protocol (https://www.fda.gov/media/134922/download). In brief, 20 *µ*L reactions included 8.5 *µ*L of Nuclease-free Water, 1.5 *µ*L of Primer and Probe mix (IDT, 10006713), 5 *µ*L of TaqPath 1-Step RT-qPCR Master Mix (ThermoFisher, A15299) and 5 *µ*L of the template. Nuclease-free water was used as negative template control (NTC). Amplification was performed using following program: 25C for 2 min, 50C for 15 min, 95C for 2 min followed by 45 cycles of 95C for 3 s and 55C for 30 s. Run data was analyzed in SDS software v1.4 (Applied Biosystems). The NTC showed no amplification throughout the 40 cycles of qPCR.

#### RT-PCR and SARS-CoV-2 genome sequencing

For sequencing 10 *µ*L of RNA from SARS-CoV-2 positive patient sample was reverse transcribed using SuperScript IV (Thermo Fisher Scientific) according to the supplier’s protocol. The protocol developed by ARTIC Network was used to generate and sequence amplicon library that covers whole SARS-CoV-2 genome on Oxford Nanopore using ligation sequencing kit (SQK-LSK109) (https://artic.network/ncov-2019) (Grubaugh et al., 2019; Tyson et al., 2020). Briefly, two multiplex PCR reactions were performed with primer pools from ARTIC nCoV-2019 V3 Panel (IDT, Table S3) using Q5 High-Fidelity DNA Polymerase (New England Biolabs). Reactions were performed with the following thermocycling conditions: 98C for 2min, 30 cycles of 98C for 15 s and 65C for 5 min. Two resulting amplicon pools for each patient sample were combined and used for library preparation. After end repair (NEB E7546) samples were barcoded using Native Barcoding Expansion Kits EXP-NBD104 and EXP-NBD114 from Oxford Nanopore. A total of 24 barcoded samples were pooled together and Nanopore adaptors were ligated. After clean-up 20 ng of multiplexed library DNA was loaded onto the MinION flowcell for sequencing. A total of 0.3 Gb of raw sequencing data was collected per patient sample.

#### SARS-CoV-2 genome assembly

MinKNOW software was used to basecall raw Nanopore reads in high-accuracy mode. ARTIC bioinformatic pipeline for COVID-19 was used to analyze reads (https://artic.network/ncov-2019). Pipeline included demultiplexing with guppy barcoder and generating consensus sequences with minimap2 and calling single nucleotide variants with nanopolish relative to Wuhan-Hu-1/2019 reference genome (Li, 2018; Quick et al., 2016; Wu et al., 2020). Consensus sequences were uploaded to GISAID (https://www.gisaid.org/), accession IDs are provided in Supplemental Table S1.

#### RT-PCR and Sanger sequencing

To verify the ORF7a^Δ115^ mutation we used RT-PCR with primers that flank the deletion region (Table S3). Reactions were performed with Q5 High-Fidelity DNA Polymerase in 25 *µ*L volume (New England Biolabs) using following program: 98C for 2 min, 35 cycles of 98C for 15 s and 65C for 5 min, 35 cycles. PCR products were analyzed on 1% agarose gels stained with SYBR Safe (Thermo Fisher Scientific). The remaining volume of the reaction was purified using DNA Clean & Concentrator kit (Zymo Research) and sent to Psomagen for Sanger sequencing. Each PCR product was sequenced with both forward and reverse primers used for PCR. To detect sgRNAs of ORF7a and ORF7b primers annealing to Leader region of SARS-CoV-2 and inside the ORF were used (Table S3). PCR products were cut out from 1% agarose gel and purified with Zymoclean Gel DNA Recovery Kit. Purified products were sequenced with forward and reverse primers. Sequence was verified by aligning to SARS-CoV-2 genome in UGENE software (Unipro).

#### Phylogenetic and ORF7a mutational analysis

An alignment of 181,003 SARS-CoV-2 genomes was downloaded from the GISAID database at 9:11 AM on 2020-11-11, and sequences without corresponding metadata entries were removed. The nucleotide positions encoding ORF7a in the reference sequence (Wuhan-Hu-1; EP ISL 402125) were extracted for the remaining 180,971 sequences, and ORF7a sequences with 100% sequence identity were clustered using CD-HIT (with settings:-c 1 -aL 1). One representative was randomly selected from each of the 846 ORF7a sequence clusters and mutations in each nucleotide and translated sequence were determined using the Biostrings package in R. Graphs of these mutations were rendered with the ggplot2 package. To construct a phylogenetic tree of global and Bozeman SARS-CoV-2 sequences, up to ten sequences from each country in the world were selected for each month of the pandemic using the Filter utility in the Augur pipeline (–group-by country year month – sequences-per-group 10). This resulted in an alignment of 4,235 GISAID sequences. Then, the NextClade online tool was used to determine the quality of 73 SARS-CoV-2 genomes (% genome covered) sequenced from Bozeman patients. 56 Bozeman sequences that were determined to be of “Good” quality were merged with the alignment of GISAID sequences, as well as an alignment of 669 representative SARS-CoV-2 genomes that had non-synonymous mutations in ORF7a. The bat coronavirus RaTG13 (MN996532.2) and the Wuhan-Hu-1 SARS-CoV-2 sequences were also merged, resulting in an alignment of 4,959 full genome sequences, and positions that are hypervariable or prone to sequencing artifacts were masked with a VCF file downloaded from EMBL at 12:45PM on 2020-05-29. The alignment was trimmed of columns composed of 90% or more gaps using Trimal (-gt 0.9), and IqTree was used to construct a maximum likelihood phylogeny (-m GTR -ninit 2 -n 2 -me 0.05). Branch lengths and internal nodes were rescaled on the resulting tree using TreeTime, and tree was re-rooted to the RaTG13 sequence using the APE package before being visualized with the ggTree package in R.

#### Western Blot

293T-hACE2 cells were transfected using Lipofectamine 3000 (Invitrogen, L3000-015) with plasmids encoding for ORF7aWT, ORF7a^Δ115^ or control pEGFP-N1 plasmid. To detect ORF7b expression cells were infected with ORF7aWT or ORF7a^Δ115^ SARS-CoV-2 strains at MOI = 0.05. After 24 hours, cells were washed two times with PBS and lysed in RIPA buffer (150 mM sodium chloride, 1% NP-40, 0.5% sodium deoxycholate, 0.2% SDS, 50 mM Tris, pH 8.0) at 4C for 30 min. The lysates were clarified by centrifugation (10,000 g, 20 min) and stored at −80C. For Western blot lysates were mixed with 6xLaemmli SDS-PAGE buffer and heated at 98C for 5 min, resolved in 12% SDS-PAGE gel and transferred onto a PVDF (ORF7b) or nitrocellulose (ORF7a) membrane using Mini Trans-Blot Electrophoretic Transfer Cell (Bio-Rad, #1703930). Membranes were blocked and probed with indicated antibodies. The proteins were visualized using Pierce ECL Western Blotting Substrate (ThermoFisher Scientific, #32106) and exposed to X-Ray film (sc-201696, Santa Cruz Biotech).

#### Immunocytochemistry

HEK 293T-hACE2 were transfected with plasmid DNA using Lipofectamine 3000 (Invitrogen REF: L3000-015) according to manufacturer’s instructions. Cells were either transfected with expression vectors for SARS-CoV-2 ORF7aWT, SARS-CoV-2 ORF7a^Δ115^, or mock transfected. 24 hours after transfection, cells were detached and seeded at 50% confluency on glass coverslips (Fisherbrand, #12-545-80) pre-coated with poly-D-lysine (Cultrex, #3439-100-01) in a 24 well plate. The next day, media was removed, and cells were washed with PBS and fixed with ice-cold 100% methanol on ice for 10 minutes. Half of the methanol was replaced with PBS three times before the methanol solution was discarded and cells were washed three times in PBS (5 minutes each wash). Cells were blocked with 1% BSA (Fisher Scientific BP9703-100) in PBS + 0.1% Tween-20 (PBST) for 30 minutes at room temperature. After blocking, cells were stained with mouse monoclonal anti-Flag antibody (Invitrogen, MA1-91878) (1:200 in 1% BSA in PBST) overnight at 4C. The next day, cells were washed 3X in PBS (5 min each) and stained with secondary antibody diluted 1:2000 in 1% BSA in PBST for 1 h at RT in the dark (Goat anti-mouse AlexaFluor488; Invitrogen, A11001). After 3x PBS washes nuclei were stained with Hoechst 33342 in PBS for 5 minutes at RT. Coverslips were then mounted with ProLong Gold antifade mounting media (Invitrogen, P36934) on Superfrost Plus microscope slides (#22-037-246). Immunostained cells were imaged using a Leica SP8 confocal microscope.

#### SARS-CoV-2 replication assays

HEK 293T-hACE2 and Vero E6 cells were seeded in the 48-well plate and infected with ORF7aWT or ORF7a^Δ115^ SARS-CoV-2 strains at MOI = 0.05 in 50 *µ*L of the cell culture media. Infections were performed in triplicates. Next, 0.5 h post infection (hpi) 500 *µ*L of the fresh cell culture media was added to each well. The supernatants (140 ul) from the infected cells were harvested at the following time points 0.5, 6, 24, 48, 72 and 120 hpi. The RNAs from the supernatants were extracted using QIAamp Viral RNA Mini Kit (QIAGEN) and used as an input in RT-qPCR with CDC N1 and N2 primers and probes as described above. Relative quantification was used (ΔCt method) to calculate viral replication versus 0.5 hpi timepoint as 2-ΔCt. The infected cells were harvested 0.5, 6 and 24 hpi and washed with PBS. Total RNA from cells was extracted using RNeasy kit (QIAGEN, Cat No./ID: 74104) with on-column DNase digestion step (QIAGEN, 79254) according to the manufacturers protocol. Viral RNA was quantified using RT-qPCR CDC N1 primers and probe. Human ACTB endogenous control was quantified with TaqMan assay (ThermoFisher, 4333762T). ΔΔCt method was used to normalize viral RNA level to host RNA and 0.5 hpi. Viral replication was calculated for each replicate as 2-ΔΔCt.

#### Type I interferon response assay

HEK 293T-hACE2 were seeded in the 48-well plate and infected with ORF7aWT or ORF7a^Δ115^ SARS-CoV-2 strains at MOI = 0.05 in 50 *µ*l of the media. After 30 min on infection the 500 *µ*L of the cell culture media was added to each well. At 24 hpi the cells were washed once with PBS, detached using Trypsin and the total RNA was extracted using RNeasy kit (QIAGEN, Cat No./ID: 74104) with on-column DNase digestion (QIAGEN, Cat No./ID: 79254) according to the manufacturer’s protocol. Non-infected cells were processed using the same protocol with mock infection. After extraction, 0,5-1 *µ*g of total RNA was reverse transcribed Reverse using SuperScript IV Reverse Transcriptase (Thermo Fisher Scientific) according to the supplier’s protocol. The cDNA was diluted 10-fold in water and analyzed with Type I interferon response (SAB Target List) H96 qPCR array. Each reaction contained 2 *µ*L of cDNA, 10 *µ*L of 2X SsoAdvanced Universal SYBR Green Supermix (Bio-Rad) and 8 *µ*L of water. Reactions were performed on QuantStudio 3 Real-Time PCR System instrument with following thermocycling conditions: 95C for 2min, 40 cycles of 95C for 5 s and 60C for 30 s. After amplification, melt curve analysis was performed to examine product specificity. Assay was performed in triplicates for non-infected cells and both viruses. Run data was analyzed in PrimePCR Analysis software (Bio-Rad). Expression of IFN-I response genes was normalized to geometric mean of 3 reference transcripts (TBP, GAPDH, HPRT1) and non-infected control (ΔΔCt method).

#### QUANTIFICATION AND STATISTICAL ANALYSIS

All statistical analyses were performed in RStudio v1.2.1335. Moving averages (n = 7) for symptom onset and positive COVID-19 tests data were calculated using geom ma function from tidyquant package and plotted with ggplot2 in RStudio. RT-qPCR data is shown as mean of three biological replicates (each with three technical replicates) *±* standard deviation (SD). Replication of viruses and expression of IFN-I response genes was compared using Student’s t-test. Medians of IFN-I response between two viruses were compared with Wilcoxon signed-rank test. Significance levels: * p *<*0.05, ** p *<*0.01, *** p *<*0.001 or ns (no significant difference, p *>*0.05).

## Data Availability

SARS-CoV-2 genome consensus sequences generated in the study were uploaded to GISAID (https://www.gisaid.org/), accession IDs are provided in Supplemental Table S1

## Acknowledgements

We are grateful to members of Bozeman Health that provided de-identified patient samples. Specifically, C. Nero, D. Smoot, W. Wallace, C. Faurot-Daniels, V. Lawrence and M. Blauvelt. We are also grateful to M. Flenniken, K. Daughenbaugh, and other members of the COVID task force at MSU for assistance establishing the COVID testing center. We thank the Lefcort lab for generous use of their fluorescent confocal microscope and Dr. Pincus for providing the HEK 293T-hACE2 cells. Research in the Wiedenheft lab is supported by the National Institutes of Health (1R35GM134867), the Montana State University Agricultural Experimental Station, the MJ Murdock Charitable Trust, the Gianforte Foundation and the MSU Office of the Vice President for Research. We thank the GISAID’s EpiFlu− Database and contributing laboratories (Table S4). The phylogenetic analysis in this paper would not have been possible without their willingness to share data.

## Author contributions

Conceptualization, B.W., A. Nemudryi, and A. Nemudraia; Methodology, B.W., A. Nemudraia, A. Nemudryi, D.S and J.H.; Sample acquisition, D.B. and B.W.; Investigation & Data Collection, A. Nemudraia., A. Nemudryi, D.S., J.H., J.N., H.L., K.V.; Genomics and bioinformatic analysis, T.W., A. Nemudryi, C.C.; Writing – Original Draft, B.W., A. Nemudryi, A. Nemudraia. and T.W.; Writing – Review & Editing, B.W, A. Nemudryi, A. Nemudraia, T.W., M.J.

