## Supplementary Figures for "SARS-CoV-2 genomic surveillance identifies naturally occurring truncations of ORF7a that limit immune suppression"

### Supplemental Data

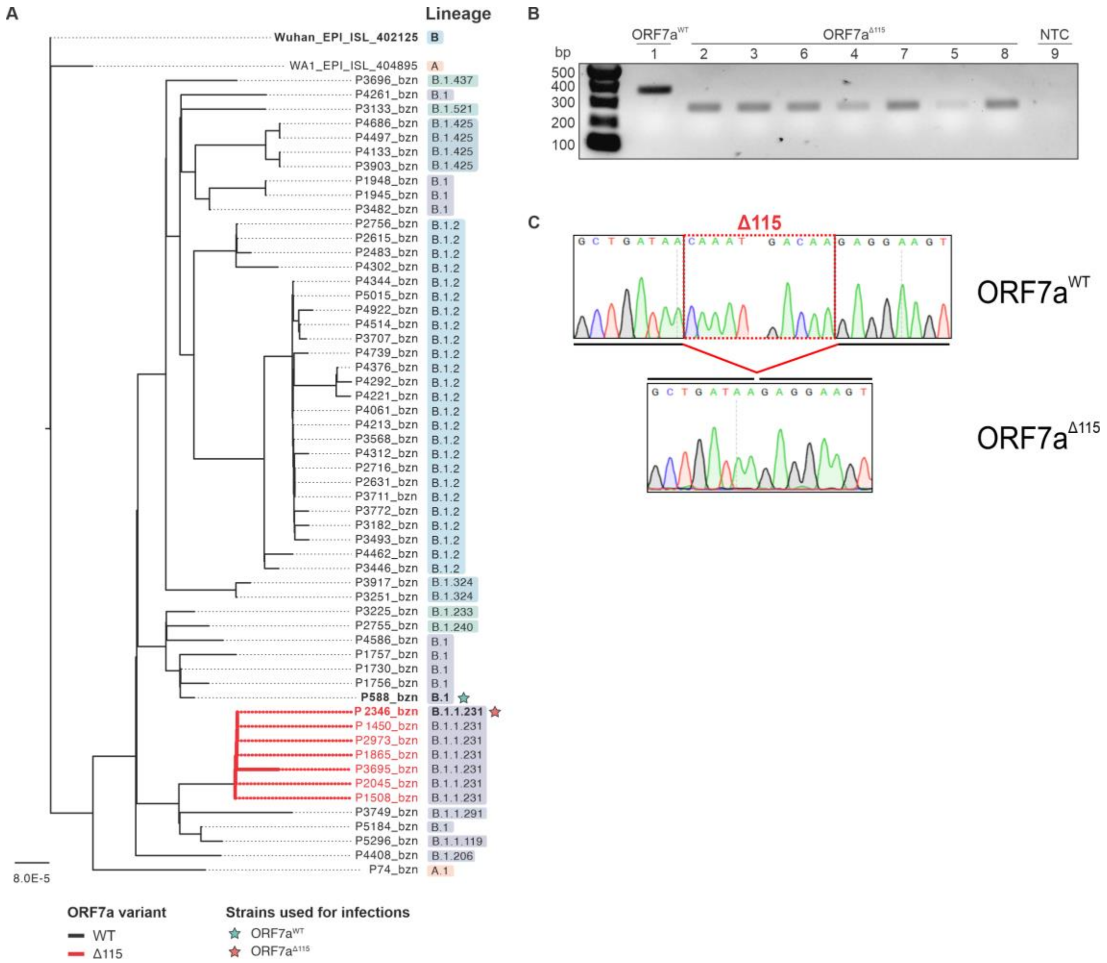


**Supplemental Figure S1, Phylogenetic and mutational analysis of ORF7a gene sequences in Bozeman SARS-CoV-2 isolates. Related to main Figure 1. A)** A maximum-likelihood phylogenetic tree indicates that ORF7a$^{\Delta115}$ strains form a monophyletic clade (red branches). Viral isolate P588 (teal star) was chosen as a "wildtype" control in our experiments. This strain shares a recent common ancestor with the predominant ORF7a$^{\Delta115}$ clade. SARS-CoV-2 lineages were assigned using pangolin utility (cov-lineages.org, (Rambaut, Andrew et al., 2020)). **B)** RT-PCR with primers targeting regions that flank the ORF7a deletion. **C)** All PCR products in (**B**) were Sanger sequenced. Representative sequencing chromatograms are shown**.** Deleted region is shown with dotted red box.

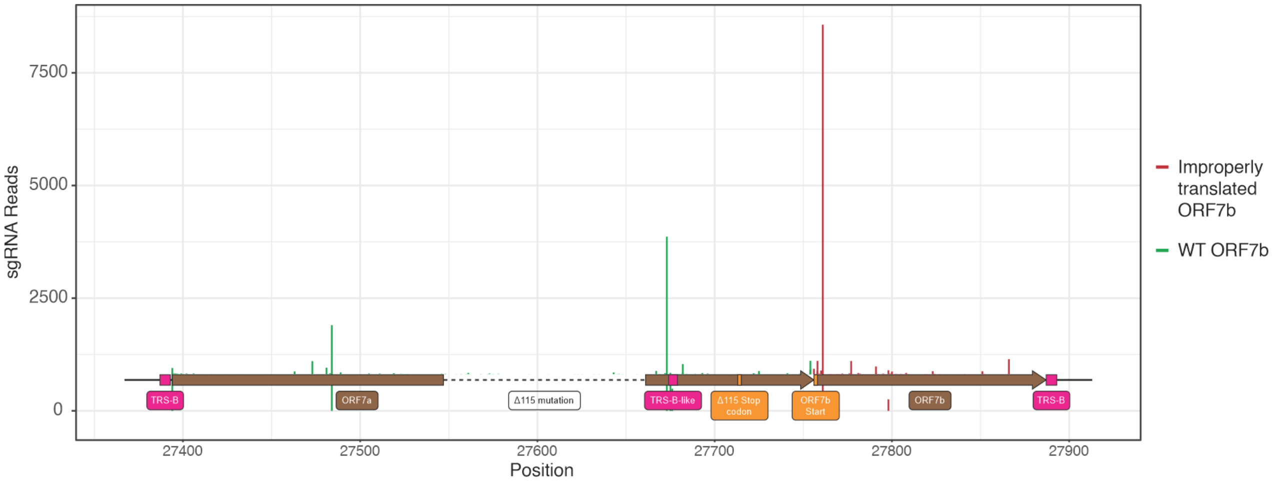


**Supplemental Figure S2, Mapping TRS-B sites required for ORF7a and ORF7b discontinuous sgRNA transcription. Related to Main Figure 2F-H.** Direct RNA sequencing data published by Kim et al was used to map TRS-B sites in SARS-CoV-2 ORF7a and ORF7b genes (Kim et al., 2020). Vertical lines indicate number of reads spanning the junction between TRS-L and “body” of the gene. Dashed line indicate region deleted in ORF7a$^{\Delta 115}$ strain. The deletion does not remove any of the mapped TRS-B sites suggesting that it has not affected ORF7b sgRNA transcription.

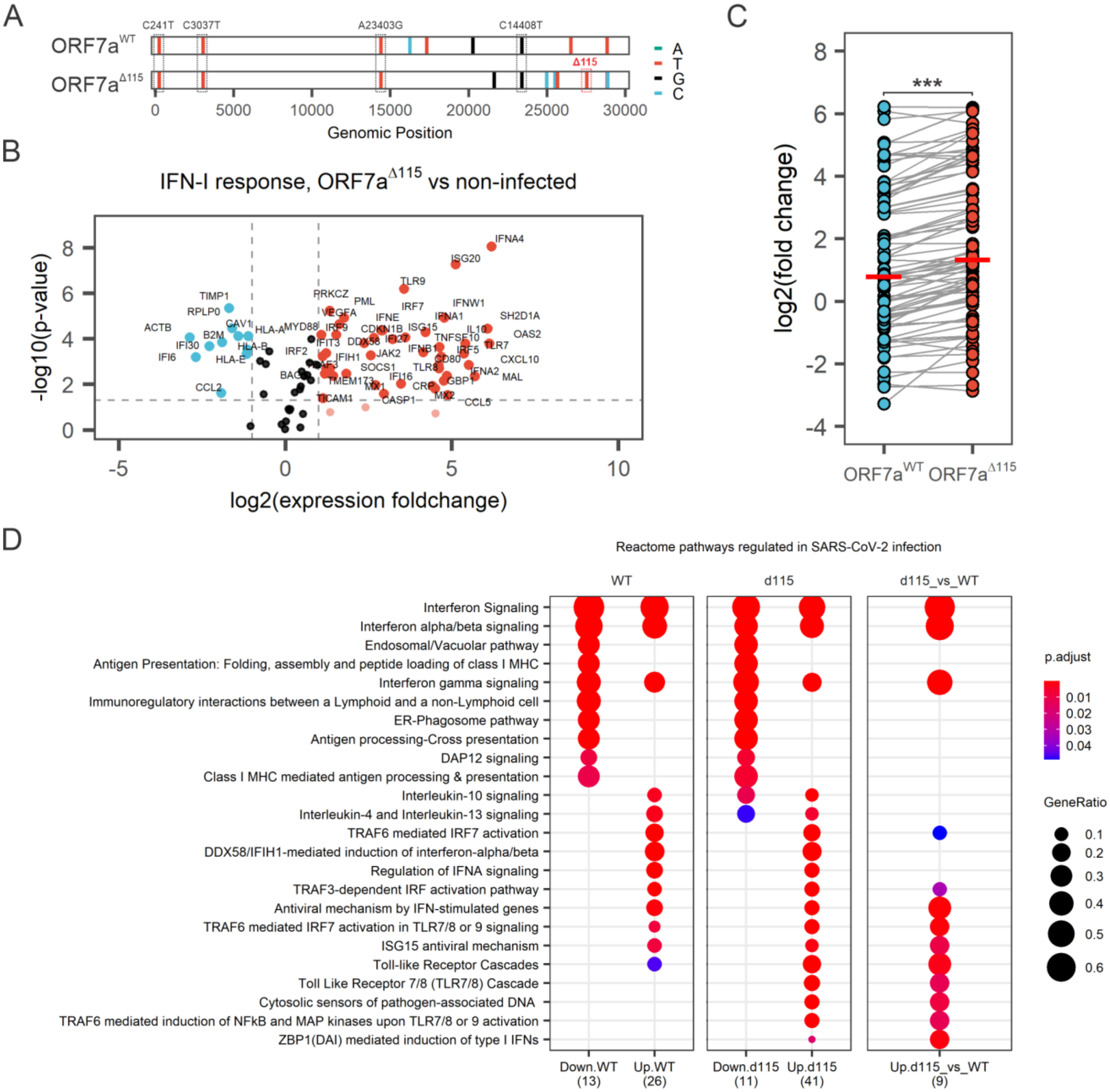
A
ORF7aWT
ORF7aΔ115
C241T
C3037T
A23403G
C14408T
Δ115
A
T
G
C
0
5000
10000
15000
20000
25000
30000
Genomic Position
B
IFN-I response, ORF7aΔ115 vs non-infected
-log10(p-value)
log2(expression foldchange)
IFNA4
ISG20
TLR9
PRKCZ
PML
IRF7
IFNW1
TIMP1
RPLP0
VEGFA
IFNE
IFNA1
SH2D1A
ACTB
CAV1
MYD88
IRF9
CDKN1B
ISG15
IL10
OAS2
B2M
HLA-A
IFIT3
DDX58
IFI27
TNFSF10
TLR7
IFI30
HLA-B
IRF2
IFNB1
IRF5
IFI6
HLA-E
IFIH1
JAK2
CD80
CXCL10
SOCS1
TLR8
IFNA2
MAL
TMEM173
MX1
IFI16
GBP1
CCL2
CRP
MX2
TICAM1
CASP1
CCL5
-5
0
5
10
C
log2(fold change)
***
ORF7aWT
ORF7aΔ115
D
Reactome pathways regulated in SARS-CoV-2 infection
WT
d115
d115_vs_WT
Interferon Signaling
Interferon alpha/beta signaling
Endosomal/Vacuolar pathway
Antigen Presentation: Folding, assembly and peptide loading of class I MHC
Interferon gamma signaling
Immunoregulatory interactions between a Lymphoid and a non-Lymphoid cell
ER-Phagosome pathway
Antigen processing-Cross presentation
DAP12 signaling
Class I MHC mediated antigen processing & presentation
Interleukin-10 signaling
Interleukin-4 and Interleukin-13 signaling
TRAF6 mediated IRF7 activation
DDX58/IFIH1-mediated induction of interferon-alpha/beta
Regulation of IFNA signaling
TRAF3-dependent IRF activation pathway
Antiviral mechanism by IFN-stimulated genes
TRAF6 mediated IRF7 activation in TLR7/8 or 9 signaling
ISG15 antiviral mechanism
Toll-like Receptor Cascades
Toll Like Receptor 7/8 (TLR7/8) Cascade
Cytosolic sensors of pathogen-associated DNA
TRAF6 mediated induction of NFkB and MAP kinases upon TLR7/8 or 9 activation
ZBP1(DAI) mediated induction of type I IFNs
Down.WT (13)
Up.WT (26)
Down.d115 (11)
Up.d115 (41)
Up.d115_vs_WT (9)
p.adjust
0.01
0.02
0.03
0.04
GeneRatio
0.1
0.2
0.3
0.4
0.5
0.6

**Supplemental Figure S3, Related to Main Figure 3. IFN-I response and gene set enrichment analysis. A)** Genome sequences of ORF7a$^{WT}$ and ORF7a$^{\Delta 115}$ are compared to Wuhan-Hu-1 reference sequence. Colored lines mark differences to reference genome. Mutations that define haplotype of B.1 SARS-CoV-2 lineage are boxed. **B)** Volcano plot depicting IFN-I response in HEK 293T-hACE2 cells infected with ORF7a$^{\Delta 115}$ SARS-CoV-2 strain at MOI = 0.05. RNA was extracted from infected cells 24 hpi and reverse transcribed. Expression of IFN-I response genes was studied using RT-qPCR array targeting 91 human transcripts (88 targets and 3 references). Experiment was performed in 3 biological replicates. Dashed lines show regulation (≥2-fold) and statistical significance thresholds (p-value < 0.05). Each dot represents mean (n = 3) normalized expression of a single gene relative to non-infected host. Genes that passed the threshold are labeled. **C)** Plot showing distribution of log2(expression fold change) values in ORF7a$^{WT}$ and ORF7a$^{\Delta 115}$ infection vs non-infected control. Each dot represents relative normalized expression for one transcript targeted with the RT-qPCR array. Gray lines connect values for the same transcript in the infection with ORF7a$^{WT}$ and ORF7a$^{\Delta 115}$ variants. Horizontal red bar shows median value. Median ISG activation between two infections was compared using Wilcoxon signed-rank test (***, p-value < 0.001) **D)** List of IFN-I response genes that were significantly regulated (p-value < 0.05, ≥ 2-fold change in expression) upon infection with ORF7a$^{\Delta 115}$ and ORF7a$^{WT}$ (vs non-infected control) were analyzed for enrichment of associated Reactome Pathways (Jassal et al., 2019; Zhou, Z. et al., 2020). Gene set enrichment analysis (GSEA) was performed using clusterProfiler package in RStudio (Yu and He, 2016; Yu et al., 2012). *Down.WT* – genes downregulated during infection with ORF7a$^{WT}$; *Up.WT* – genes upregulated during infection with ORF7a$^{WT}$; *Down.d115* – Reactome pathways enriched in genes downregulated during infection with ORF7a$^{\Delta 115}$; *Up.d115* – genes upregulated during infection with ORF7a$^{\Delta 115}$. *Up.d115_vs_WT* – Reactome pathways enriched in genes upregulated during infection with ORF7a$^{\Delta 115}$ vs ORF7a$^{WT}$ (**Figure 3E**).
